# SARS-CoV-2 antibodies detected in human breast milk post-vaccination

**DOI:** 10.1101/2021.02.23.21252328

**Authors:** Jill K. Baird, Shawn M. Jensen, Walter J. Urba, Bernard A. Fox, Jason R. Baird

**Author notes:** Corresponding author: Jason R. Baird, Earle A. Chiles Research Institute, Providence Portland Medical Center, 4805 NE Glisan St. North Pavilion, Suite 2N130G, Portland, OR 97213.

## Abstract

**Importance:** The SARS-CoV-2 pandemic has infected over a hundred million people worldwide, with almost 2.5 million deaths at the date of this publication. In the United States, Pfizer-BioNTech and Moderna vaccines were first administered to the public starting in December 2020, and no lactating women were included in the initial trials of safety/efficacy. Research on SARS-CoV-2 vaccination in lactating women and the potential transmission of passive immunity to the infant through breast milk is needed to guide patients, clinicians and policy makers during the worldwide effort to curb the spread of this virus.

**Objective:** To determine whether SARS-CoV-2 specific immunoglobins are found in breast milk post-vaccination, and to characterize the time course and types of immunoglobulins present.

**Design:** Prospective cohort study

**Setting:** Providence Portland Medical Center, Oregon, USA

**Participants:** Six lactating women who planned to receive both doses of the Pfizer-BioNTech or Moderna vaccine between December 2020 and January 2021. Breast milk samples were collected pre-vaccination and at 11 additional timepoints, with last sample at 14 days post 2^nd^ dose of vaccine.

**Exposure:** Two doses of Pfizer-BioNTech or Moderna SARS-CoV-2 vaccine.

**Main Outcome(s) and Measure(s):** Levels of SARS-CoV-2 specific IgA and IgG immunoglobulins in breast milk.

**Results:** In this cohort of 6 lactating women who received 2 doses of SARS-CoV-2 vaccine, we observed significantly elevated levels of SARS-CoV-2 specific IgG and IgA antibodies in breast milk beginning at Day 7 after the initial vaccine dose, with an IgG-dominant response.

**Conclusions and Relevance:** We are the first to show that maternal vaccination results in SARS-CoV-2 specific immunoglobulins in breast milk that may be protective for infants.

## INTRODUCTION

The severe acute respiratory syndrome coronavirus 2 (SARS-CoV-2) pandemic has infected over 111 million people worldwide to date, causing nearly 2.5 million deaths. ^1^ Two novel mRNA vaccines encoding the viral spike protein have recently been FDA-approved and administered to patients in the United States beginning December 2020, with potential to slow rate of infection and decrease incidence of severe cases that lead to hospitalization and death. The initial trials for the Pfizer-BioNTech BNT162b2 and Moderna mRNA-1273 vaccines excluded pregnant and breastfeeding women, ^2, 3^ and there is currently a dearth of evidence to guide women, clinicians, and policy makers on whether to recommend vaccination for these populations.

Previous research has shown that women infected with SARS-CoV-2 produce antibodies in their breast milk against the virus. While one group found a robust secretory-IgA dominant response in human milk from 15 women after infection with SARS-CoV-2, ^4^ other researchers showed SARS-CoV-2 specific secretory IgA in milk samples collected both pre-pandemic (2018) and during Spring 2020. ^5^ While findings from that study were limited by lack of confirmed SARS-CoV-2 infection in any of the subjects who donated milk, the authors suggest that secretory IgA found in breast milk has a flexible binding pocket which makes it polyreactive to a range of pathogens, while IgG seemed to be more specific for prior SARS-CoV-2 infection, with an increase in SARS-CoV-2 specific IgG in the 2020 group and in milk from subjects who reported recent respiratory illness.

To date, there has been no published research on the breast milk antibody response to the SARS-CoV-2 vaccine. Our objective in this report is to confirm the presence of these immunoglobulins and more specifically to characterize the typology and time course of the antibody response in breast milk after vaccination. We hypothesize that we will detect significantly elevated IgA and IgG anti-SARS-CoV-2 spike protein antibodies in breast milk relative to pre-vaccine levels within 14 days post-vaccination.

## METHODS

### Participants

Breast milk samples were collected from 6 women between December 2020 and February 2021. All women were living in Oregon, currently breastfeeding, and were already scheduled to receive the SARS-CoV-2 vaccine at time of recruitment. There were no known prior exposures to COVID-19 at the time of recruitment. Sample collection was requested at the following timepoints: pre-vaccination, 1, 4, 7, 11, and 14 days post-1^st^ vaccine dose, 1 day before 2^nd^ dose, and 1, 4, 7, 11, and 14 days post-2^nd^ vaccine dose (booster). Samples were dated and accepted if collected within 24 hours of the requested timepoint. Written consent to use milk samples for research was obtained. Sample collection was approved by the institutional review board (IRB #06-108A) of Providence Health & Services.

### Sample collection and preparation

Human milk samples (2-4 mL) were collected at home with clean electric breast pumps into sterile plastic containers and stored immediately at - 20° C. Samples were kept frozen and transported on ice in coolers to Providence Portland Medical Center laboratory, where they were stored at − 80□°C in an industrial freezer. The samples (*n*□= 6) were rapidly thawed at 37□°C in a water bath and centrifuged at 1301□×□*g* for 20□min at 4□°C. After removing the fat layer, the supernatant was collected, separated into aliquots and stored at − 80□°C until analysis.

### Anti-spike ELISA

96-well NUNC MAXISORP ELISA plates were coated with 1.0µg/ml of SARS-CoV2 Spike protein prefusion-stabilized ectodomain (LakePharma, #46328) overnight at 4° C. The following day plates were washed with PBS PBS/0.5% Tween and blocked with PBS/0.5%Tween containing 5% Blotting-Grade Blocker (Bio-Rad, #170640404) for 2 hrs at 37° C. Plates were washed, and titrated milk samples were aliquoted and incubated for 1 hr at room temperature. Plates were washed with PBS/0.5% Tween, followed by incubation with either anti-IgG HRP (1:10000, FisherScientific, #A18811) or anti-IgA HRP (1:8000, Jackson ImmunoResearch, #109-035-011) for 30 min at room temperature. Plates were washed with PBS/0.5% Tween and exposed to SureBlue TMB Peroxidase Substrate (VWR, #95059-286) for 45 minutes. The TMB reaction was stopped with 1M Phosphoric Acid, and the optical absorbance (450, 1.0 sec) for each well was read on a Wallac Victor Microplate Reader. To determine the relative concentration of volunteer Spike-specific IgG or IgA antibodies, protein dilutions of IgG or IgA (250ng/ml, 125ng/ml, 62.5ng/ml, 31.25ng/ml, 15.6ng/ml, 7.8 ng/ml, 3.9ng/ml, and none) were coated in the first column of wells on each ELISA plate. This titration curve was incubated with the same anti-IgG HRP (1:10000, FisherScientific, #A18811) or anti-IgA HRP (1:8000, Jackson ImmunoResearch, #109-035-011) as the rest of the ELISA plate as described above. A titration curve equation was determined by simple linear regression of the OD 450 readings. Volunteer sample concentrations were determined from the titration curve equation multiplied by the dilution factor and expressed at U/mL (1U/mL = 1ng/ml of IgG or IgA protein standard).

## RESULTS

We recruited six lactating women with no known prior SARS-CoV-2 infection who received 2 doses of the Pfizer-BioNTech BNT162b2 (3 subjects) or Moderna mRNA-1273 (3 subjects) vaccine. All six women received both doses of vaccine and completed the study. We received baseline (pre-vaccine) timepoint samples from all women with the exception of Subject 4. In total, 50 human milk samples were included in the analysis (7-9 timepoints per subject).

### Presence and levels of SARS-CoV-2 spike IgG and IgA in breast milk

We found significantly elevated levels of anti-spike IgG and IgA relative to pre-vaccine baseline (Figure 1) and characterized individual levels and kinetics of anti-SARS-CoV-2 spike protein IgG (Figure 2) and IgA (Figure 3) production in breast milk post-vaccination. None of our subjects had significant levels of SARS-CoV-2 specific immunoglobulins in breast milk prior to vaccination. For all subjects, levels of virus-specific IgG and IgA increased at Day 7 post-vaccine, and this was consistent between Moderna and Pfizer-BioNTech vaccine. Levels of IgG and IgA broadly decreased prior to the administration of the 2^nd^ vaccine dose, and sharply increased in the timepoints after the booster. There was significant variability between subjects in the magnitude and time course of the antibody response, but no detectable difference in levels of antibody produced between the two vaccines.

**Figure 1:**
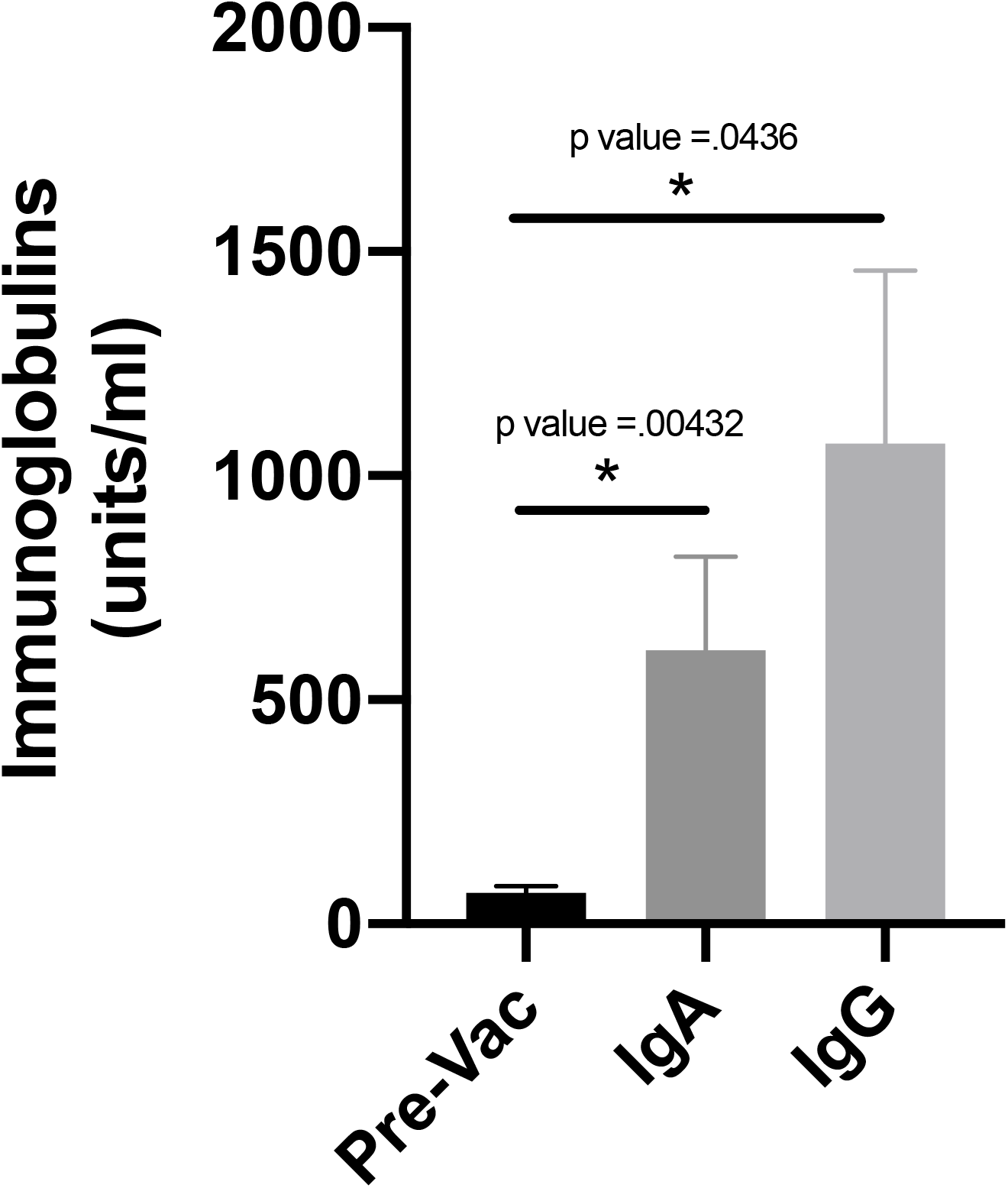
Histogram of pre- and post- vaccination (Day 11 post-boost) levels (in ng/mL) of anti-SARS-CoV-2 spike IgG and IgA in breast milk (n=5).

**Figure 2:**
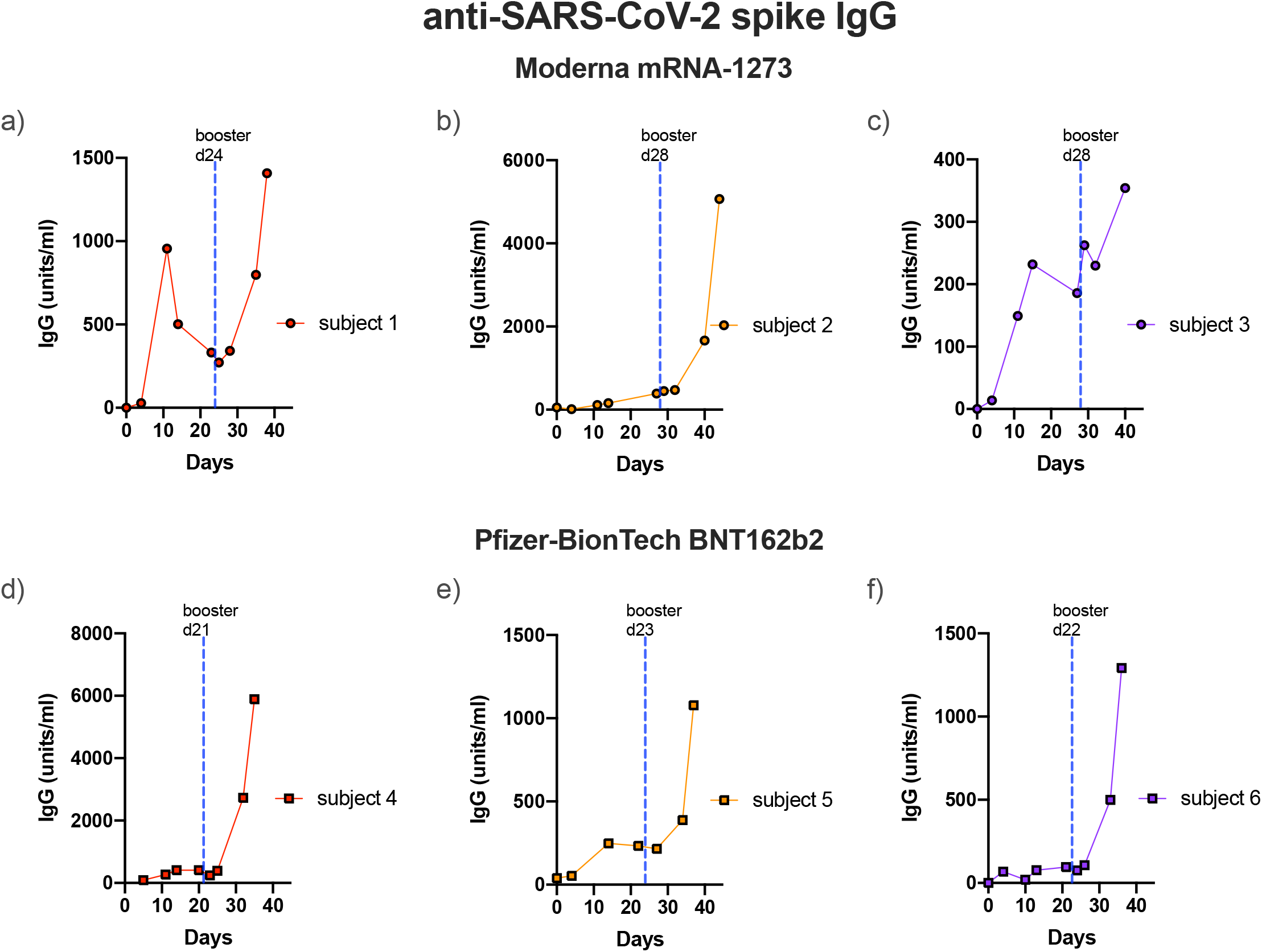
Levels (in ng/mL) and kinetics of anti-SARS-CoV-2 spike IgG in breast milk post- vaccination. Subjects 1 (a), 2 (b) and 3 (c) received Moderna vaccine, while subjects 4 (d), 5 (e), and 6 (f) received Pfizer-BioNTech vaccine. Dotted line represents date of 2^nd^ dose (booster) administration for each subject.

**Figure 3:**
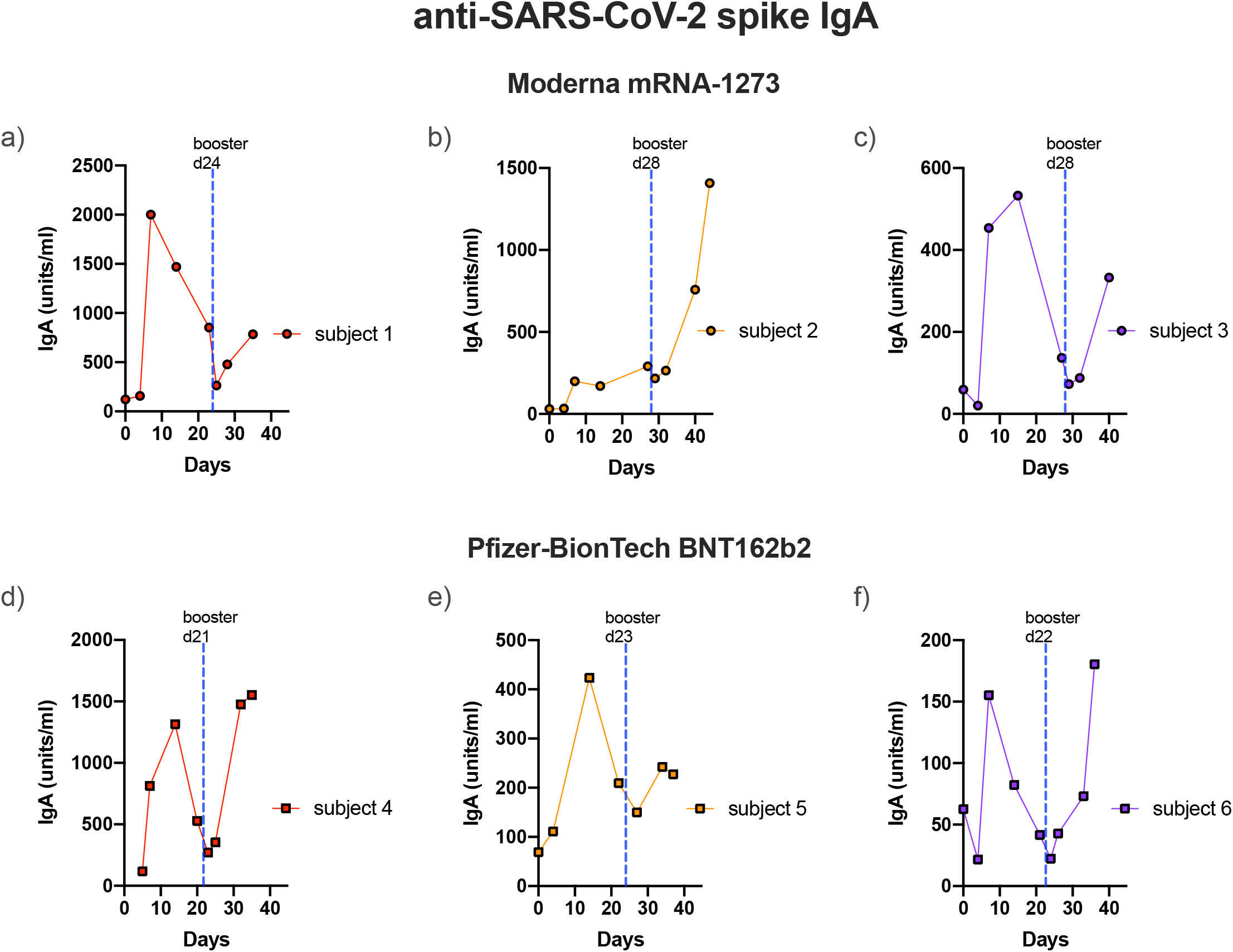
Levels (in ng/mL) and kinetics of anti-SARS-CoV-2 spike IgA in breast milk post- vaccination. Subjects 1 (a), 2 (b) and 3 (c) received Moderna vaccine, while subjects 4 (d), 5 (e), and 6 (f) received Pfizer-BioNTech vaccine. Dotted line represents date of 2^nd^ dose (booster) administration for each subject.

## DISCUSSION

Since the first confirmed case of COVID-19 in November 2019, the SARS-CoV-2 virus has spread rapidly worldwide. The toll of this pandemic has been especially devastating in the United States, with 28.2 million confirmed cases and over 500,000 deaths to date. ^1^ There are two SARS-CoV-2 vaccines currently approved for use in the US, with no research to date on their use in breastfeeding women, a group excluded from the initial clinical trials. In this paper, we analyzed breast milk samples from a cohort of 6 women who received 2 doses of either the Pfizer-BioNTech or Moderna vaccine and found a significant increase in IgA and IgG immunoglobulins specific to the viral spike protein starting between day 7 and day 14 post-vaccination. This time course is consistent with previous findings of the immunogenicity of these two vaccines. In the Phase I trial of Moderna mRNA-1273 vaccine, increased anti-spike serum IgG was seen at day 15.^6^ In preliminary analysis of BNT162b2 (Pfizer-BioNTech), IgG increase to convalescent-equivalent levels was seen at Day 21, while at Day 8 serum IgG was still at or near baseline levels.^7^

In contrast to previous work showing an IgA-dominant antibody response in the breast milk of previously infected/exposed women,^4^ our results indicate that the post-vaccine antibody response to SARS-CoV-2 in breast milk is IgG dominant. One potential explanation for this difference is that vaccinated mothers are exposed to viral antigen via intramuscular injection whereas infected mothers have immunological education occurring in the mucosa, where IgA plays a greater role.

While establishment of a breast milk antibody response to the SARS-CoV-2 vaccine is novel and promising, further research is needed on the longevity of the antibody response in breast milk as well as the magnitude and duration of effect on infant immunity to the virus. More imminently, further characterization of the breast milk immune response including IgG antibody subtypes, IgM levels, and spike protein blocking assay in our upcoming studies will flesh out this picture.

## CONCLUSION

Currently, there is little to no research to guide lactating women and their healthcare providers when deciding whether or not to get vaccinated. We provide the first evidence that mothers vaccinated against SARS-CoV-2 produce antibodies to this virus in breast milk that may be protective for infants.

## Data Availability

Raw data were generated at Earle A. Chiles Research Institute. Derived data supporting the findings of this study are available from the corresponding author JRB on request

## FUNDING

This work was supported by generous grants from Nancy Lematta (BAF) and the Chiles Foundation (BAF).

## Notes

### Competing Interest Statement

The authors have declared no competing interest.

### Author Declarations

Providence health and services IRB

### Summary of Updates

The only thing we changed was the corresponding author email address.

